# Propylene Oxide in Exhaled Breath as a Marker for Discriminating TMAU-like Conditions from TMAU

**DOI:** 10.1101/2024.04.11.24305677

**Authors:** Irene S. Gabashvili

## Abstract

Volatile Organic Compounds (VOCs) triggering respiratory irritation are implicated in conditions such as Trimethylaminuria (TMAU) and “people are allergic to me” (PATM) which occur in otherwise healthy individuals without clear syndromic associations. Despite the absence of established non-targeted non-challenge-based diagnostic procedures, recent studies have identified discriminatory VOCs associated with PATM using gas chromatography-mass spectrometry. Breath VOCs, originating from the bloodstream, hold promise for non-invasive diagnosis.

We conducted breath analysis on 23 individuals exhibiting TMAU-like symptoms and identified a diverse array of volatile organic compounds (VOCs) that discriminate between different subgroups. Using logistic regression, we achieved an accuracy of 88%, with both precision and recall at 88-89%, in distinguishing TMAU-negative individuals from those who tested positive at some point in their lives, solely based on the presence of Propylene Oxide ((2R)-2-Methyloxirane and (2S)-2-Methyloxirane). However, due to the limited subset and missing data, it cannot serve as the sole discriminating biomarker. Inclusion of additional VOCs in the analysis increased accuracy of the model to 85-95%, with precision and recall ranging from 85% to 100%, depending on the combinations of VOCs used.

Unsupervised learning algorithms generally grouped positively tested individuals together based on endogenous VOCs, while negatively tested individuals were clustered into two distinct groups. Toluene, previously found to be elevated in PATM individuals, was identified as a discriminatory marker for those previously diagnosed with TMAU, but had since tested negative while still experiencing symptoms. Other PATM biomarkers, such as p-Xylene and Hexanal, were generally higher in TMAU positive individuals and were good predictors of TMAU history when combined with other VOCs.

Our analysis revealed that the TMAU-positive group exhibited a greater abundance of biomarkers indicative of advanced oxidative stress in their breath samples and primary oxidative stress in their air samples, likely originating from their skin. Conversely, the TMAU-negative group demonstrated a higher likelihood of biomarkers associated with secondary oxidative stress in their air samples.

Our findings highlight the potential of breath analysis as a non-invasive diagnostic tool for idiopathic malodor conditions. They underscore the importance of analyzing exogenous chemicals for insights into metabolism, detoxification, and elimination of toxins. This approach could help eliminate unnecessary challenge tests and emphasize the role of metabolomics in understanding the mechanisms underlying these conditions.

**Trial registration:** ClinicalTrials.gov NCT03451994;

https://clinicaltrials.gov/study/NCT03451994

**Data:** available at https://osf.io/w7682 via the Open Science Framework

## 1. Introduction

The concept that breath contains molecules indicative of normal or abnormal physiological states traces back to the dawn of medicine. Technological advancements have significantly enhanced breath analysis, rendering it more effective in diagnosing and monitoring health conditions. This improvement stems from its ability to detect a diverse array of volatile organic compounds (VOCs), which arise from environmental exposures, cellular metabolism, oxidative stress, and microbiome activity. VOCs serve as biomarkers for a spectrum of diseases including asthma, chronic obstructive pulmonary disease, cystic fibrosis, tuberculosis, liver disease, lung cancer, and COVID-19, as well as for individual traits such as identity, sex, age, and circadian rhythms [1].

Idiopathic malodor conditions, which involve the excretion of volatile organic compounds (VOCs) like trimethylamine (TMA), are among the conditions that could significantly benefit from VOC analysis due to their association with distinct odors. Additionally, the phenomenon of People Are Allergic To Me (PATM), where individuals might emit non-odorous but potentially irritating VOCs like toluene and xylene [2], highlights the complex interplay between exogenous and endogenous compounds in breath.

Non-syndromic idiopathic odor conditions, though complex and less appealing to funded researchers, have prompted studies driven by sufferers for over 15 years. Conditions coined by sufferers, including MEBO (Metabolic body odor), IMBS (intestinal metabolic bromhidrosis syndrome), Bloodborne body odor and halitosis, Systemic odor, and PATM, have spurred extensive questionnaire surveys, blood and urine test donations, and microbiome profiling. Successful tests, such as choline and glucose challenge tests, have revealed similarities among these conditions and the medically recognized trimethylaminuria (TMAU), correlating with symptom severity levels [3]. However, notable differences persist, particularly in response to environmental stimuli like diet [4,5], rendering each sufferer unique and necessitating further data for comprehensive understanding.

This study aims to explore the breath VOC profiles of individuals with TMAU and related conditions, assessing the potential of both endogenous and exogenous compounds as disease biomarkers. By understanding the relationship between breath VOCs and these conditions, we aim to contribute to the advancement of non-invasive diagnostic tools and deepen our comprehension of the metabolic foundations of idiopathic malodor conditions.

## 2. Methods

### 2.1 Human Subjects

This study utilized a cross-sectional design and included participants who met the predetermined inclusion and exclusion criteria. The protocol for participant selection and data collection was previously established and published [6]. In 2012-2013, participants attended one in-person data collection session in four cities: Chicago, Illinois; Miami, Florida; New York, New York (USA) and London (UK).

The following procedures were conducted:

- Informed Consent: Participants provided informed consent prior to any study-related procedures.
- Medical Records Review: Relevant medical records were reviewed to collect pertinent clinical information.
- Breath Sample Collection

The participants were also asked to fill a baseline survey and use Aurametrix to record their daily food intake, activities and symptoms. Some of the participants participated in other studies, before and after breath study, and gave consent to use their data for analysis.

### 2.2 Data Collection

Breath samples were collected and concentrated using the portable Breath Collection Apparatus (BCA) 5.0, originally developed by Michael Phillips and refined over time [6,7]. This device, controlled by a microprocessor, collects alveolar breath onto an adsorbent tube, with collection duration and flow rate settings adjustable on the device. VOCs were captured onto sorbent traps for subsequent analysis via automated thermal desorption, gas chromatography, and mass spectrometry.

All collections were conducted in the morning following an overnight fast. Participants wore a nose clip and breathed for two minutes through a disposable valved sterile mouthpiece unit and a bacterial filter into the breath collection apparatus. Separate sorbent traps were used for collecting breath samples from alveolar breath and room air, which were then sealed for laboratory analysis. To ensure accurate alveolar gradient calculations, all subjects had been breathing room air for at least one hour in the same environment as the breath collection.

VOCs were thermally desorbed from sorbent traps and assayed via gas chromatography-mass spectrometry (GC-MS) with the addition of an internal standard (1-bromo-4-fluorobenzene) for normalization and chromatogram alignment [8]. This internal standard was included with every chromatographic assay of both breath and air samples to quantify peak areas and control for instrument drift.

### 2.3 Data Analysis

Data from each chromatographic peak, including chemical identity and area under the curve (AUC) were used for further analysis. The alveolar gradient for each VOC was calculated as (AUC VOC in breath/AUC internal standard) – (AUC VOC in air/AUC internal standard).

The analysis involved logistic regression to investigate the separation between two groups based on selected variables, controlling for confounders such as dairy intake, age, gender, and choline. The logistic regression model’s performance was evaluated using metrics like precision, recall, F1-score, and accuracy. Statistical analyses were conducted using Python’s statistical and machine learning libraries.

Non-parametric tests, such as the unpaired Mann-Whitney-Wilcoxon test and the Wilcoxon signed-rank test, were used for comparing groups without assuming a normal distribution. The predictive ability of various biomarkers was assessed using a logistic regression model, with its accuracy determined as the proportion of correctly classified instances.

A sensitivity analysis was conducted using bootstrap sampling techniques to ensure the robustness of the results. The logistic regression model was trained and evaluated on multiple bootstrap samples, with the mean and standard deviation of accuracy scores calculated to assess the model’s stability.

To determine the optimal number of clusters for k-means clustering, the silhouette score was used.

Outlier detection was performed using z-score normalization, with a threshold commonly set to 3 to identify data points lying more than three standard deviations from the mean. Fisher’s exact test was employed to assess the statistical significance of differences in proportions between groups for specific conditions.

## 3. Results

### 3.1 User Statistics

The study began with 25 individuals expressing interest and completing breath testing. However, due to the need for retesting, we ultimately obtained valid data from 23 participants aged 20-. All participants were followed up, and their data were analyzed. Table 1 displays the characteristics of these individuals, excluding those who dropped out after requiring retesting, having previously tested negative for trimethylaminuria (TMAU).

**Table 1.**
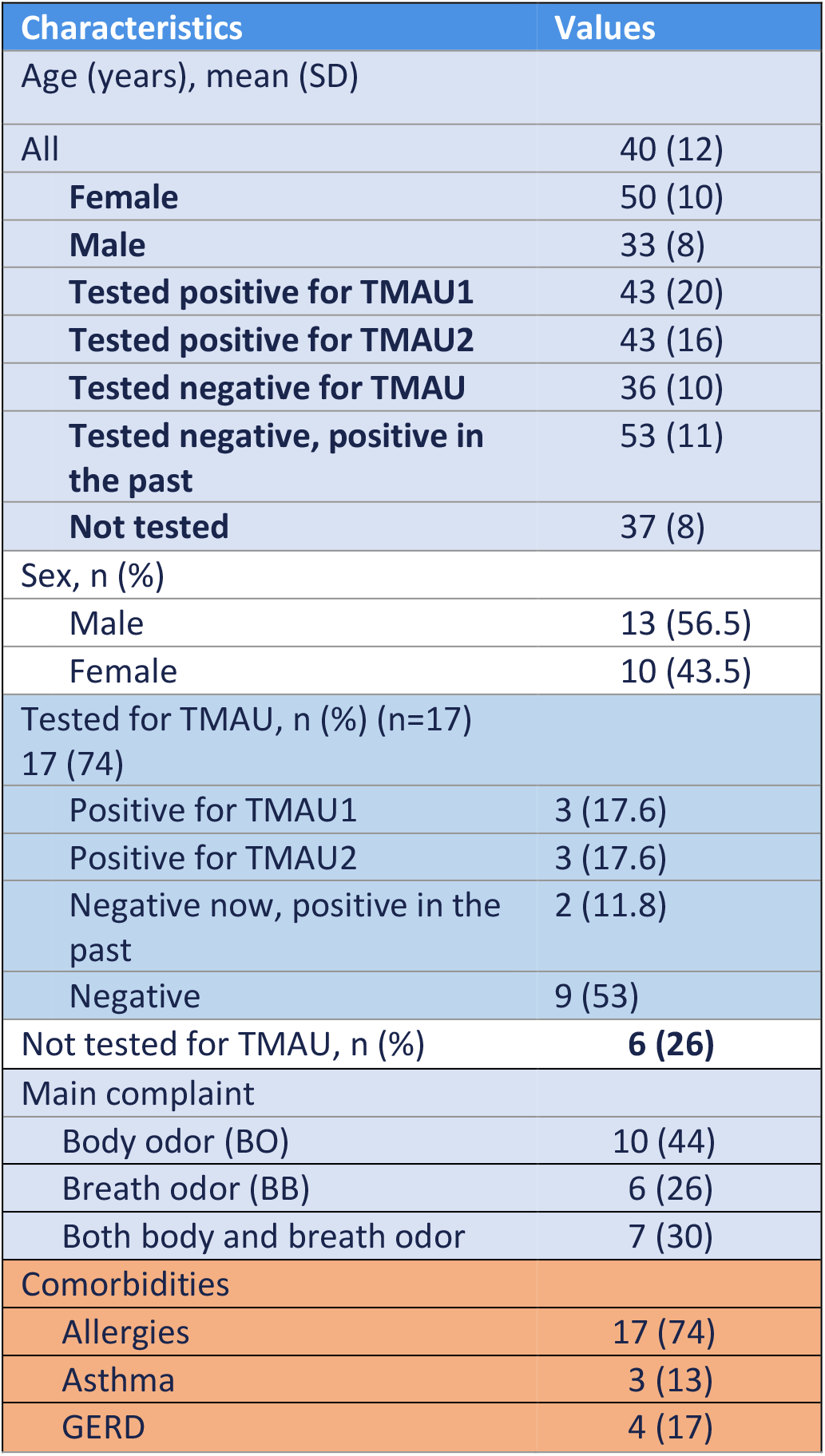
Baseline demographic and clinical characteristics of study participants (N=23, n=17 of whom previously tested for TMAU).

### 3.2 Evaluation Outcomes

The analysis of breath samples revealed a diverse array of volatile organic compounds (VOCs), with a total of 3,253 unique VOCs observed across 14,063 peaks. On average, each breath sample contained over 600 different VOCs, with a mean of 623 ± 58, a minimum of 474, and a maximum of 744. The data showed a slightly higher number of negative gradients (7,407) compared to positive gradients (6,657), indicating a greater prevalence of VOCs originating from external sources. Propylene Oxide (PO) and its S-isomer, S-methyloxirane, were the most abundant compounds in most samples, particularly in individuals who tested negative for Trimethylaminuria. Some but not all of them were smokers.

Figure 1 illustrates the abundance of Propylene Oxide or its S-isomer, S-methyloxirane, in the exhaled breath of study participants compared to the abundance in the air. Green squares represent TMAU-negative individuals, red rhombuses are TMAU1 individuals (as diagnosed by urine test), yellow circles represent TMAU2, blue squares with stars inside represent individuals who tested positive for TMAU1 or TMAU2 earlier in their lives but their most recent test was negative. Gray triangles are those who never tested for TMAU.

**Figure 1.**
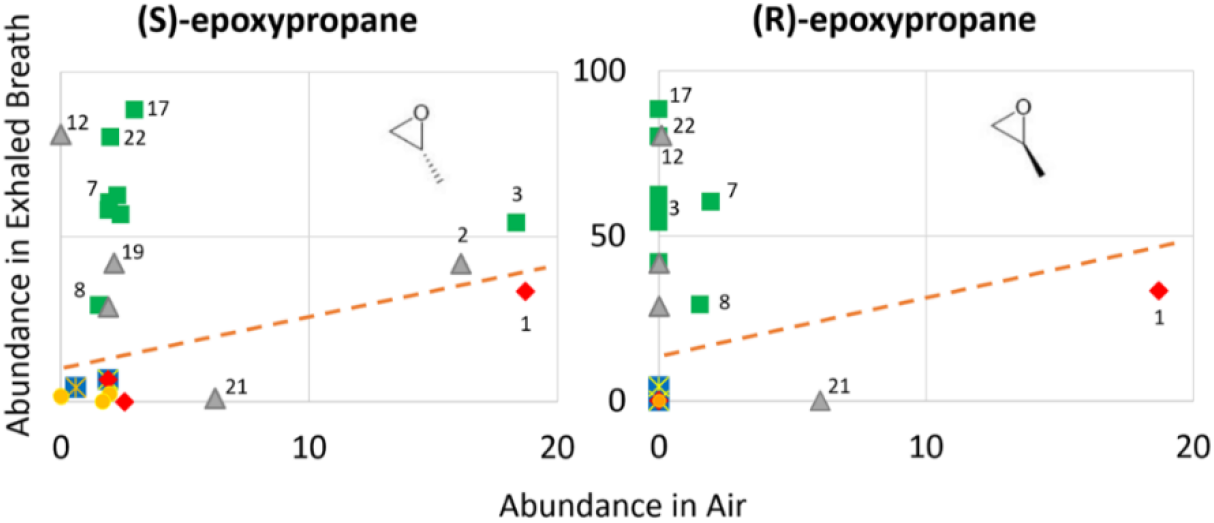
Comparison of Propylene Oxide Abundance in Exhaled Breath Versus Ambient Air among Study Participants

R-isomers of Propylene Oxide were not detected in the air or breath of eight individuals, and two of these individuals did not have S-isomers in their breath. Gradients were negative only for TMAU-positive individuals.

Given the small subset and missing data, Propylene Oxide alone cannot be solely relied upon as the discriminating biomarker between TMAU-positive and negative individuals. Our analysis included all other VOCs present in the samples, and we applied various prefiltering approaches and unsupervised and supervised machine learning algorithms.

Based on z-score normalization with a threshold of 3, which identifies points more than 3 standard deviations away from the mean as outliers, it appears that every sample in the dataset has at least one feature considered an outlier. This illustrates the need for personalized approaches, given the wide variation in feature values, many unique points, and heavy-tailed distributions of values.

Using k-means clustering on all or subsets of features, we observed that TMAU-positive individuals cluster better together, while negative individuals form at least two or more different clusters. We separately considered VOCs more likely to be exogenous vs. endogenous, based on whether the gradients were positive or negative. The TMAU-positive group seemed to be better clustered according to endogenous VOCs (positive gradients) but not exogenous VOCs (negative gradients), indicating a more pronounced effect on the body’s internal processes than on the interaction with external compounds. On the other hand, TMAU-negative individuals clustered about the same for both endogenous and exogenous VOCs, suggesting less variation in metabolic processes vs external interactions within this group.

Using PCA, we separated TMAU-negative and positive individuals for some combinations of features accounting for over 95% of variance. Using logistic regression without accounting for confounding variables, we achieved an accuracy range of 85% to over 95% in predicting TMAU diagnosis across various filtering scenarios. The models demonstrated a precision and recall of 85-100%, indicating their reliability in correctly identifying the target outcome. The performance remained consistent even after controlling for potential confounders such as dairy or choline intake, prolonged antibiotics use, age, and gender, underscoring the robustness of the selected variables in effectively distinguishing between different groups.

We note that VOCs typically associated with unpleasant odors (eg, Indole, Acetic Acid, Phenol) were notably underrepresented in our study cohort. These compounds did not effectively differentiate between the sub-cohorts, suggesting a potential dependency on younger age (late stages of puberty) rather than the history of TMAU testing.

A scatter plot (Figure 2) shows a composite score calculated using a combination of five features (Hydroperoxide, hexyl; Hexanal; Decane, 2-methyl-; Tetradecane; Decane, 2,6,6-trimethyl-) with their respective coefficients. The alveolar gradient of Propylene Oxide is plotted on the y-axis. Higher X values for TMAU-positive individuals (right side of the plot) signify that they were more likely to have oxidative stress biomarkers in their breath and more likely to exhale VOCs known as biomarkers of asthma and COPD [9-12], even though the majority did not have any of these conditions. Additionally, we noted elevated levels of certain volatile organic compounds (VOCs) associated with conditions such as celiac disease, Crohn’s disease, ulcerative colitis, nonalcoholic fatty liver disease [11], and liver disease (D-limonene, known to correlate with blood with bilirubin, albumin and INR, as well as hepatic disease severity, [13]). These conditions, while rare comorbidities in the MEBO community, underscore the significance of VOCs in health assessments.

**Figure 2.**
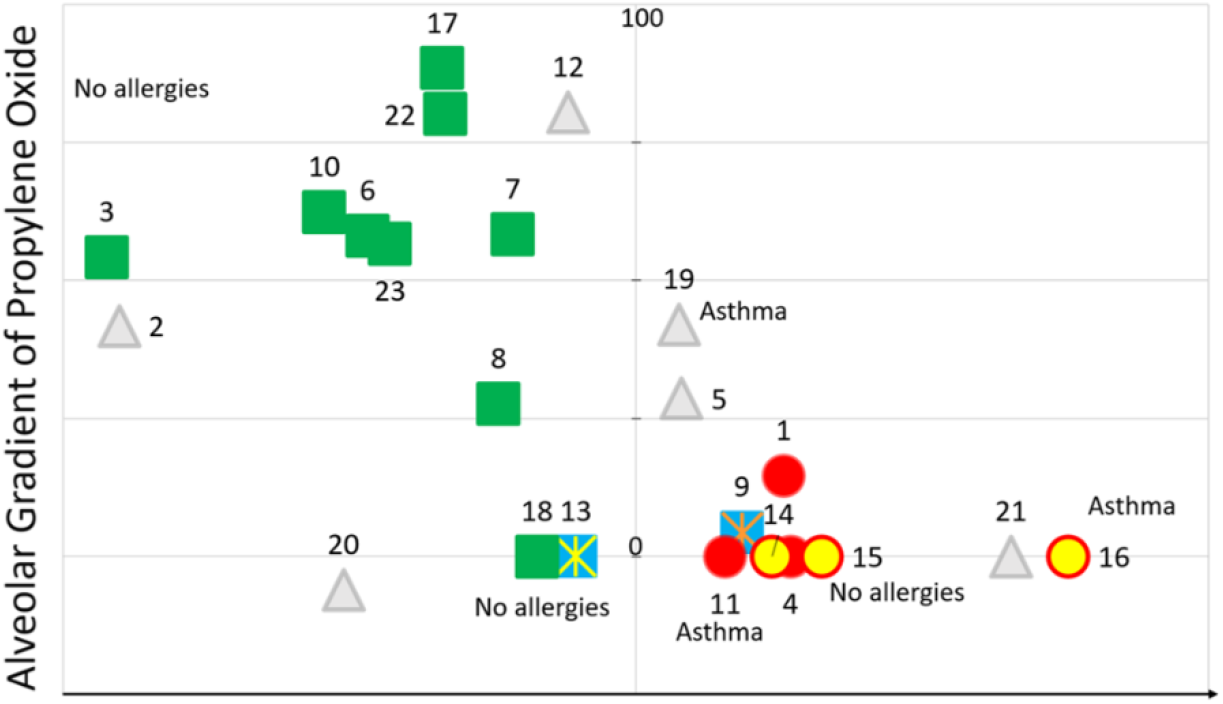
Scatter plot showing a composite score derived from five VOCs, reflecting the progression of inflammatory response, plotted against the alveolar gradient of Propylene Oxide

## Discussion

### Principal Results

Our analysis of breath samples has unveiled a diverse array of volatile organic compounds (VOCs), with a higher number of negative gradients suggesting a greater prevalence of exogenous VOCs. The notable abundance of Propylene Oxide and its S-isomer in TMAU-negative individuals suggests that these compounds could be indicative of delayed elimination, bioaccumulation/microbial biofilms trapping compounds or altered enzymatic processing, possibly due to overreactive cytochromes (eg, synthesizing PO from Propylene) or alterations in other enzymes such as GST [14]. Variants in GSTT1 and GSTM1 can affect PO detoxification, with null genotypes potentially increasing susceptibility to its toxic effects. GSTT1-/-genotype showed higher mortality in COVID-19 patients [15] while GSTP1 Ile/Val genotype was found to be associated with a higher risk of developing a severe form of COVID-19 in the population of vaccinated patients [16]. In our subsequent studies [17], we observed some notable differences between the MEBO and average populations, but the incidence of COVID-19 was significantly lower in the MEBO group compared to the average population.

The reliance on Propylene Oxide alone as a discriminating biomarker is limited due to the small subset and missing data, but this finding warrants further exploration of potential environmental sensitivities in individuals with TMAU-like conditions.

The application of machine learning algorithms has demonstrated the utility of VOCs in predicting TMAU diagnosis with high accuracy and reliability. The distinct clustering of TMAU-positive individuals based on endogenous VOCs suggests a more pronounced effect of FMO3 deficiency of on internal metabolic processes in these individuals. In contrast, TMAU-negative individuals show less variation in metabolic processes versus delayed elimination of exogenous substances, suggesting different metabolic dynamics in these groups.

Our results also suggest that TMAU-positive samples are characterized by a higher degree of oxidative stress, as indicated by elevated levels of secondary lipid peroxidation products and other biomarkers. This group exhibited a greater abundance of biomarkers indicative of advanced oxidative stress in their breath samples and primary oxidative stress in their air samples, likely originating from their skin. Conversely, the TMAU-negative group demonstrated a higher likelihood of biomarkers associated with secondary oxidative stress in their air but not breath samples.

The clear distinction between TMAU-positive and negative groups based on the composite scores highlights the potential of breath biomarkers in differentiating between the metabolic profiles of different TMAU-like conditions. The findings underscore the complexity of metabolic interactions and the potential of metabolomics in understanding the mechanisms of these conditions.

The study also points to the need for personalized approaches in analyzing breath samples, given the wide variation in feature values and heavy-tailed distributions.

### Comparison with Prior Work and Limitations

Our study’s findings align with existing research on the association of VOCs with various diseases and the potential of breath analysis as a non-invasive diagnostic tool [1,2,4,5,7-13,18]. Previous studies have highlighted the role of compounds highlighted in this study, such as D-limonene [13] and Tetradecane [11], in disease diagnosis and metabolic profiling. Our work builds upon this foundation by providing a more detailed analysis of metabolic profiles associated with TMAU and related conditions, exploring the distinct subsets of these conditions, and highlighting the role of the microbiome in VOC metabolism.

One of the novel aspects of our study is the examination of exogenous compounds inhaled from the environment, in particular, Propylene Oxide, which may serve as predictors of certain conditions. This area of research is driven by the hypothesis that individuals with certain conditions may process environmental compounds differently, leading to variations in absorption, storage, or elimination.

Metabolomics data is inherently complex due to the presence of both endogenous and exogenous compounds, whose origins are not always definitively identifiable [1,5]. The concept of “human exposomics” underscores the significance of understanding the contribution of environmental factors to a healthy human breath profile. While fasting can mitigate the detection of some exogenous compounds in breath analysis, completely eliminating features originating from consumer products is impractical and does not reflect real-world environmental exposure. In fact, using exogenous compounds has been proposed as a strategy to assess the activity of metabolic enzymes and organ function through breath analysis [18].

Our study’s focus on the exogenous VOC Propylene Oxide contributes to the advancement of environmental breath biomarker research. Despite being detected in human breath this compound has not been extensively studied to date. [14]

The study has limitations. The relatively small sample size may impact the generalizability of our findings. Additionally, while the logistic regression and Linear Discriminant Analysis (LDA) models demonstrate good predictive ability, further validation with larger cohorts is warranted to confirm their robustness and suitability for clinical applications. It is also important to acknowledge the complexity of the data due to the presence of both endogenous and exogenous compounds. Some compounds initially considered endogenous, such as those emanating from the skin, can transition into exogenous compounds. As a result, determining the definitive origins of these compounds may not always be straightforward.

## Conclusions

Our study underscores the potential of breath analysis and metabolomics in identifying biomarkers for TMAU and related conditions. The findings highlight the importance of considering oxidative stress and the microbiome’s influence in disease diagnosis and understanding disease mechanisms.

A particular area of interest is the analysis of exogenous compounds such as Propylene Oxide. This interest is driven by the hypothesis that individuals with certain conditions may process environmental compounds differently, leading to variations in absorption, storage, or elimination. The study of breath VOCs, therefore, provides a unique opportunity to explore these differences and their potential impact on health.

Further research with larger cohorts is needed to validate the predictive models and explore the clinical applications of biomarkers such as Propylene Oxide in non-invasive disease screening and monitoring.

## Data Availability

Data is available at https://osf.io/w7682 via the Open Science Framework

## Data Availability

All data are available online via The Open Science Framework

https://osf.io/w7682/

## Acknowledgements

I extend my gratitude to Michael Arenibar, the study initiator and coordinator, Maria de la Torre, the founder of MEBO Research, and all the participants who contributed to the success of this work. Additionally, we acknowledge the invaluable support of Michael Phillips and Menssana Research for their assistance in processing the samples. Study volunteers self-funded their tests, and the author of this study contributed pro bono.

## Ethics Approval and Consent to Participate

Ethical review was provided by MEBO Research Institutional Review Board. The primary prospective study was approved on February 28, 2012 (MR 2012-01) and the secondary retrospective study was approved on February 15, 2018 (20111001005MEBO). The research was conducted in accordance with the principles embodied in the Declaration of Helsinki and in accordance with local statutory requirements. All participants gave written informed consent to participate in the study.

